# Evaluating Large Language Model-Generated Brain MRI Protocols: Performance of GPT4o, o3-mini, DeepSeek-R1 and Qwen2.5-72B

**DOI:** 10.1101/2025.04.08.25325433

**Authors:** Su Hwan Kim, Severin Schramm, Lena Schmitzer, Kerem Serguen, Sebastian Ziegelmayer, Felix Busch, Alexander Komenda, Marcus R. Makowski, Lisa C. Adams, Keno K. Bressem, Claus Zimmer, Jan Kirschke, Benedikt Wiestler, Dennis Hedderich, Tom Finck, Jannis Bodden

## Abstract

**Purpose:** To evaluate the potential of LLMs to generate sequence-level brain MRI protocols.

**Methods:** A dataset of 150 brain MRI cases was derived from imaging request forms obtained from the local institution. For each case, a reference MRI protocol was established by two board-certified neuroradiologists, with discrepancies resolved through consensus. GPT-4o, o3-mini, DeepSeek-R1 and Qwen-2.5-72B were employed to generate brain MRI protocols based on the case descriptions. For each model, protocol generation was conducted under two conditions: 1) with additional in-context learning involving local standard protocols and sequence explanations (enhanced) and 2) without additional external information (base). Additionally, two radiology residents independently defined MRI protocols for a subsample of 50 cases. The frequencies of redundant sequences, total missing sequences, and missing critical sequences are reported. The sum of redundant and missing sequences (accuracy index) was defined as a comprehensive metric to evaluate LLM protocoling performance. Accuracy indices were compared between groups using paired t-tests, with false discovery rate correction applied to control for multiple testing.

**Results:** The two neuroradiologists achieved substantial inter-rater agreement (Cohen’s κ = 0.74). The lowest accuracy index and therefore superior performance, was observed with o3-mini (base: 2.65; enhanced: 1.94), followed by GPT-4o (base: 3.11; enhanced: 2.23), DeepSeek-R1 (base: 3.42; enhanced: 2.37) and Qwen-2.5-72B (base: 5.95; enhanced: 2.75). o3-mini consistently outperformed the other models with a significant margin. All four models showed highly significant performance improvements under the enhanced condition (*adj. p* < 0.001 for all models), primarily driven by a substantial reduction of redundant MRI sequences. In the subsample, the highest-performing LLM (o3-mini [enhanced]) yielded an accuracy index comparable to residents (o3-mini [enhanced]: 1.92, resident 1: 1.80, resident 2: 1.44).

**Conclusion:** Our findings demonstrate promising potential of LLMs in automating brain MRI protocoling, especially when augmented through in-context learning. o3-mini exhibited superior performance, followed by GPT-4o.

## Introduction

Magnetic resonance imaging (MRI) of the brain is a complex diagnostic examination allowing for the assessment of various neurological conditions. The process of determining the appropriate sequences for brain MRI scans based on a review of the patient’s medical history and clinical question, commonly referred to as “protocoling”, is a crucial task requiring radiologists to carefully balance clinical necessity with efficiency.

On the one hand, imaging protocols should be sufficiently comprehensive to address the clinical indication. Omission of critical MRI sequences can necessitate repeat examinations (also known as callback examinations). In fact, protocol errors were reported to be the most common reason for such callbacks (28%), followed by inadequate anatomic coverage (21%) (1). On the other hand, protocols should be limited to essential sequences to minimize scan time and healthcare costs. As the demand for MRI continues to increase (2), strategies to increase patient access to MRI and to reduce wait times are a subject of ongoing interest (3–5). In addition, optimal MRI protocols can reduce unnecessary exposure to contrast agents with potential adverse effects such as allergic reactions (6).

Institutions typically employ a set of standardized imaging protocols targeted at common clinical scenarios, such as “MR brain for brain metastasis” or “MR brain for multiple sclerosis” (7). Yet, they often prove insufficient in more complex cases, requiring individualized protocol adjustments. Importantly, protocoling remains a time-consuming, non-interpretative task intensifying radiologist workload, exacerbating already increasing demands on their time (8,9). In a study analysing the workflow in an academic neuroradiology reading room, protocoling was found to take up 6.2% of the workday of radiologists, and was identified as a frequent source of interruption from image interpretation (10).

In light of these challenges, previous studies have evaluated the potential of artificial intelligence (AI) tools to select or determine radiological imaging procedures (7,11–15). Wong et al. trained a recurrent neural network to classify brain MRI cases into one out of eight predefined brain MRI protocols (7). Several studies used large language models (LLMs) to predict the most suitable imaging modality, the anatomical region, and the need for contrast application (12–15). Suzuki et al. analysed the performance of GPT-4 (Generative Pre-Trained Transformer 4) in suggesting a single brain MRI sequence to be added to a standard brain MRI protocol based on a disease name only, which limited the realism of the scenario (11). Therefore, this study aimed to evaluate the ability of LLMs to suggest granular, sequence-level brain MRI protocols based on realistic clinical cases.

## Methods

### Study Design

Ethical approval was waived by the institutional review board.

### Case Selection

A set of 150 fictitious brain MRI cases in German was developed based on real patient cases from the local imaging database. For this purpose, the condensed medical history and clinical question indicated in the imaging request form as well as demographic information was obtained. Subsequently, the original data was modified in each case by shifting the age range by 5 – 10 years and substituting the location of the primary clinical or imaging finding in the medical history with a plausible alternative (e.g. “left frontal intracerebral hemorrhage” instead of “right parietal intracerebral hemorrhage”). All references to locations were manually removed, and specific dates were replaced with relative time descriptors (e.g. “2 years ago”) to reach full anonymization. Cases were classified into five categories (vascular, neoplasia, inflammation, degenerative, miscellaneous) based on the main clinical question. In addition, cases were categorized as ‘typical’ if the reference protocol was identical to any of the local standard MRI protocols, and as ‘atypical’ if it was not.

### Brain MRI Reference Protocols

Two board-certified neuroradiologists with 7 years of experience each (JB and TF) independently defined a suitable brain MRI protocol for each of the 150 cases (Figure 1). Moreover, the two radiologists determined the critical MRI sequences per case, defined as those without which a brain MRI examination would have to be repeated, due to their clinical relevance. Cases with inter-rater disagreement were adjudicated through consensus. The level of inter-rater agreement between the two board-certified neuroradiologists on a sequence level was determined using Cohens’ kappa.

**Figure 1:**
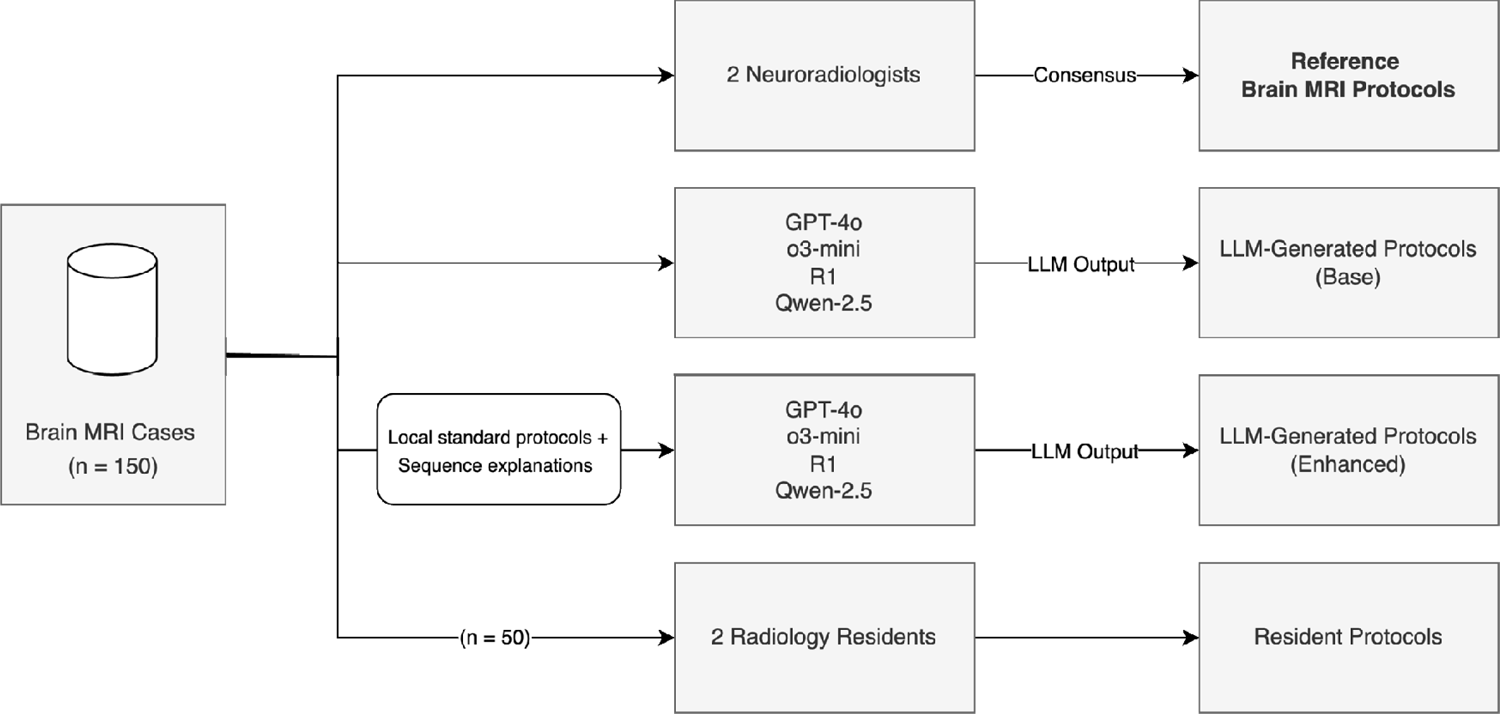
Study Design. Two board-certified neuroradiologists established the reference brain MRI protocols, with discrepancies resolved through consensus. GPT-4o, o3-mini, DeepSeek-R1 and Qwen-2.5-72B were employed to generate brain MRI protocols based on the case descriptions. For each model, protocol generation was conducted under two conditions: 1) with additional in-context learning involving local standard protocols and sequence explanations (enhanced) and 2) without additional external information (base). Additionally, two radiology residents defined MRI protocols for a subsample of 50 cases.

### LLM Selection and Access

To include state-of-the-art models from both open-weight and closed-weight categories, the following state-of-the-art LLMs were selected: GPT-4o (OpenAI, Inc., San Francisco, USA; closed-weight), o3-mini (OpenAI, Inc., San Francisco, USA; closed-weight), Qwen2.5-72B (Alibaba Cloud, Singapore; open-weight), and DeepSeek-R1 (Hangzhou DeepSeek Artificial Intelligence Basic Technology Research Co., Ltd., Hangzhou, China; open-weight).

GPT-4o and o3-mini were accessed via OpenAI’s official application programming interface (API) at https://platform.openai.com/docs/models. DeepSeek-R1 and Qwen2.5-72B were accessed via Fireworks AI, a generative AI inference platform with servers deployed in the United States and Europe (https://fireworks.ai/models). For all models, the structured output mode was applied to obtain responses adhering to a structured JSON (Javascript Object Notation) schema, enabling programmatic analysis of LLM-generated protocols. For GPT-4o, DeepSeek-R1, and Qwen-2.5-72B, a temperature setting of 0 was selected to ensure deterministic outputs, whereas the temperature parameter was not supported with o3-mini. Queries with GPT-4o and o3-mini were performed on 6 Feb 2025 and 16 Feb 2025, respectively. DeepSeek-R1 and Qwen2.5-72B queries were both executed on February 6, 2025. Model details are provided in Table 1.

**Table 1:** Model details.

| Model | # Parameters | Link | API Access |
| --- | --- | --- | --- |
| gpt-4o-2024-08-06 | Unknown | https://openai.com/index/hello-gpt-4o/ | https://platform.openai.com/docs/models/gpt-4o#gpt-4o |
| o3-mini-2025-01-31 | Unknown | https://openai.com/index/openai-o3-mini/ | https://platform.openai.com/docs/models/gpt-4o#o3-mini |
| DeepSeek-R1 | 671B | https://github.com/deepseek-ai/DeepSeek-R1 | https://fireworks.ai/models/fireworks/deepseek-r1 |
| Qwen2.5-72B-Instruct | 72B | https://huggingface.co/Qwen/Qwen2.5-72B-Instruct | https://fireworks.ai/models/fireworks/qwen2p5-72b-instruct |

### LLM Prompting

The base prompt was iteratively refined using five fictitious patient cases not included in the test dataset. In response to initial observations of misinterpretation of certain MRI sequence names, brief descriptions of the less common sequences were added to the base prompt. The final base prompt was defined as follows:

*“You are a senior neuroradiologist tasked with defining a brain MRI protocol for a given clinical case. Consider the patient’s demographics, medical history, and clinical question. Synthesize this information to define an MRI protocol. For each sequence, indicate ‘yes’ or ‘no’. Adhere to the data schema provided to you. Include only clinically relevant sequences, avoid redundant or unnecessary sequences. Here is a brief explanation of the more uncommon sequences. CISS (Constructive Interference in Steady State): A 3D gradient-echo sequence with high spatial resolution. CE_1st_pass_Angio (Contrast-Enhanced First-Pass Angiography): A dynamic imaging technique using gadolinium contrast to visualize blood vessels during the first pass of contrast through circulation. CSF_Drive: A specialized sequence for assessing CSF flow dynamics. CSF_PCA (Phase Contrast Angiography): Measures pulsatile CSF flow velocities and directions. T1_BB (Black Blood Imaging): Suppresses blood signal for vessel wall imaging. TRAK_4D (4D Time-Resolved MR Angiography with Keyhole): A 4D (time-resolved) MR angiography technique that captures the dynamics of blood flow over time. TRANCE_4D (4D Time-Resolved Angiography using Non-Contrast Enhancement): A non-contrast-enhanced 4D MRA technique, primarily used for imaging vascular structures based on cardiac-triggered sequences. In the field ‘reasoning’, indicate your rationale for each sequence included.”*

For each model, queries were executed using ‘base’ queries (without additional context) and ‘enhanced’ queries employing in-context learning, a method guiding model behavior by embedding task-relevant information in the prompt (16). In the enhanced condition, models were provided with 20 standardized brain MRI protocols from the local institution (Supplement 1), along with an explanation of the clinical indications for each of the 27 available MRI sequences (Supplement 2, created by SS and SHK), all included within the model’s context window.

One such explanation is provided in the following:

*“TOF-MRA (Time-of-Flight Magnetic Resonance Angiography) is a non-contrast, static technique used to image the intracranial arteries. It confirms the patency and course of major vessels by demonstrating normal flow, and it excludes significant stenoses or occlusions when normal flow patterns are observed. TOF-MRA is a basic sequence for static vessel imaging and should be included in protocols that aim to assess cerebral arteries.”*

Our Python code for executing LLM queries is publicly available in our GitHub repository at https://github.com/shk03/llm_brain_mri_protocols.

### Evaluation of LLM Accuracy

LLM performance in defining brain MRI protocols was evaluated by calculating the number of redundant sequences, total missing sequences, and missing critical sequences, as compared to the reference protocol. The sum of redundant and missing sequences was defined as the ‘accuracy index’ serving as an overall metric for protocoling accuracy. Furthermore, the number of cases with unnecessary contrast application (protocols with at least one post-contrast sequence) was calculated.

For the purpose of the analysis, ‘T2*’ and ‘SWI’ (susceptibility-weighted imaging) sequences were considered equivalent. Similarly, ‘T1 Dixon’ and ‘T1 MPRAGE’ were treated as interchangeable.

### Resident-Generated Brain MRI Protocols

In a randomly selected subsample of 50 out of 150 brain MRI protocols, two radiology residents further defined MRI protocols, serving as a comparison for evaluating LLM-generated protocols.

For this subsample, two radiology residents with 18 and 16 months of dedicated neuroradiology experience (KS and LS) formulated brain MRI protocols. To simulate a realistic scenario, residents were allowed to access the local standard protocols if necessary.

### Statistical Analysis

All statistical analyses were conducted in Python (version 3.13.2) using ‘SciPy’ and ‘statsmodels’ libraries. Two-sided paired t-tests were used to compare groups based on the accuracy index and the frequency of cases involving unnecessary contrast administration.

To account for the increased risk of Type I errors due to multiple testing, the Benjamini– Hochberg procedure was applied to control the false discovery rate (FDR) at 0.05. For each comparison, the FDR-adjusted p-value is reported. Statistical significance was set at p < 0.05.

## Results

### Dataset

The dataset comprised 150 cases with an equal gender distribution (50.0% female). The median patient age range was 56–60 years. ‘Vascular’ cases were most frequently observed (26.7%), followed closely by ‘Neoplasia’ cases (24.7%). Conversely, cases related to degenerative conditions were relatively underrepresented (6.7%). A majority of cases (64.7%) was categorized as ‘atypical’, based on the divergence of the respective reference protocol from any of the local standard brain MRI protocols (Table 2). The randomly selected subsample had a similar composition, as shown in Table 3.

**Table 2:** Dataset overview. 150 fictitious brain MRI cases were generated by significantly modifying real patient cases from the local imaging database. The dataset included demographic information (age range and sex), the condensed medical history, and clinical question. Number of sequences in the reference protocols are indicated as mean (± standard deviation).

| Variable | Category | Value | # Sequences in Reference Protocol |
| --- | --- | --- | --- |
| Sex | Female | 50.0% (75/150) |  |
|  | Male | 50.0% (75/150) |  |
| Median age range | - | 56 – 60 | - |
| Pathology Type | Vascular | 26.7% (40/150) | 5.4 (± 1.9) |
| | Neoplasia | 24.7% (37/150) | 5.4 (± 1.1) |
| | Miscellaneous | 23.3% (35/150) | 5.5 (± 1.3) |
| | Inflammation | 18.7% (28/150) | 6.2 (± 1.4) |
| | Degenerative | 6.7% (10/150) | 4.2 (± 0.4) |
| Protocol type | Typical | 35.3% (53/150) | 5.5 (± 1.4) |
| | Atypical | 64.7% (97/150) | 5.5 (± 1.5) |

**Table 3:**
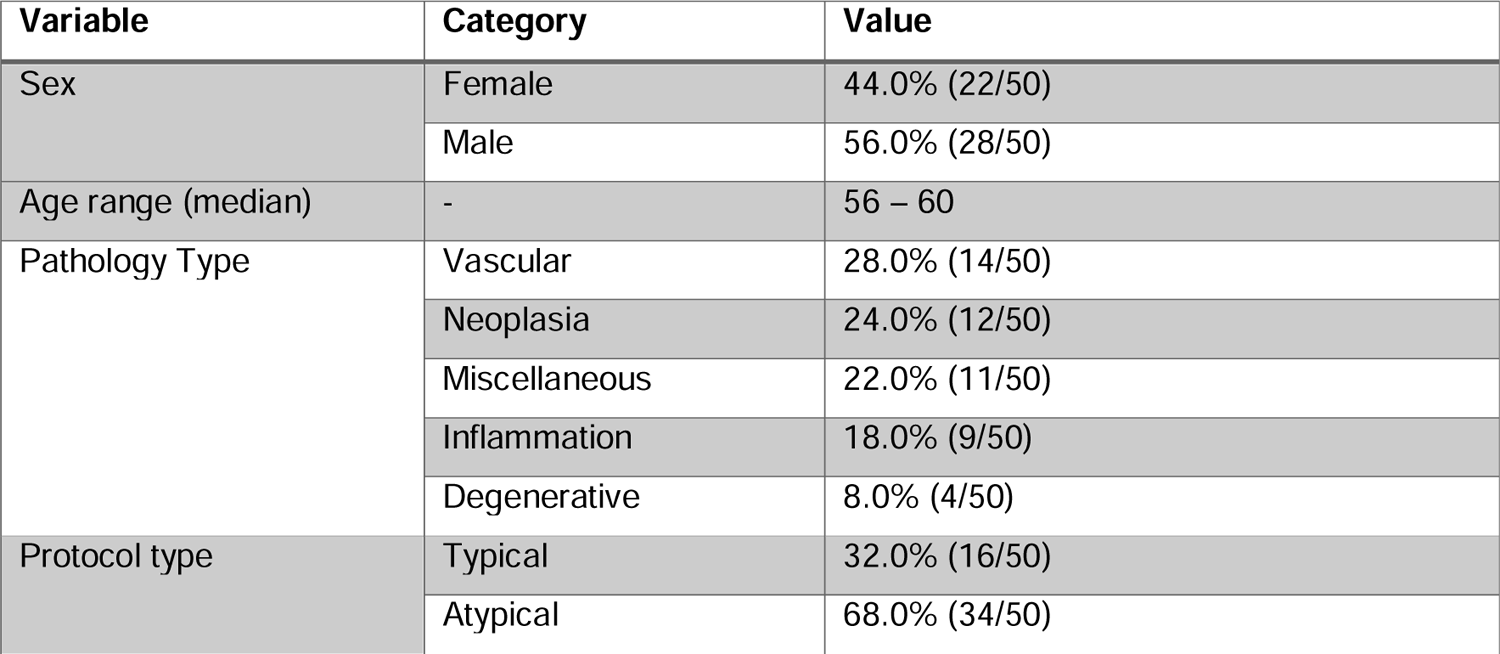
Overview of the subsample dataset (n = 50). From the original 150 brain MRI cases, a random subsample of 50 cases was selected. Two radiology residents independently defined MRI protocols for these cases, serving as a comparison for evaluating LLM-generated protocols.

| Variable | Category | Value |
| --- | --- | --- |
| Sex | Female | 44.0% (22/50) |
|  | Male | 56.0% (28/50) |
| Age range (median) | - | 56 – 60 |
| Pathology Type | Vascular | 28.0% (14/50) |
|  | Neoplasia | 24.0% (12/50) |
|  | Miscellaneous | 22.0% (11/50) |
|  | Inflammation | 18.0% (9/50) |
|  | Degenerative | 8.0% (4/50) |
| Protocol type | Typical | 32.0% (16/50) |
|  | Atypical | 68.0% (34/50) |

### Reference Protocols

The two neuroradiologists achieved substantial inter-rater agreement, reaching a concordance of 96.0% across all MRI sequences (Cohen’s κ = 0.74) and 97.3% for critical sequences (Cohen’s κ = 0.75). The reference brain MRI protocols included an average of 5.5 ± 1.50 sequences (range: 2–10), among which 3.22 ± 1.1 sequences (range: 1–6) were classified as critical. The most frequently included sequence was FLAIR (100.0%; 150/150), followed by SWI (74.0%; 111/150) and DWI (72.7%; 109/150). In comparison, TOF-MRA post-contrast, CE first-pass angiography, and dynamic post-contrast T1 sequences were infrequently employed (0.7% each; 1/150). Contrast administration was required in 65.3% (98/150) of protocols. Cases categorized as ‘inflammatory’ exhibited the highest mean sequence count (6.2 ± 1.4), whereas ‘degenerative’ cases had the lowest (4.2 ± 0.4). Yet, equal sequence counts were found in typical (5.5 ± 1.4) and atypical (5.5 ± 1.5) cases.

### LLM Protocoling Performance

A sample brain MRI case including the LLM inputs and outputs is shown in Figure 2. The lowest accuracy index, indicating superior performance, was observed with o3-mini (base: 2.65; enhanced: 1.94), followed by GPT-4o (base: 3.11; enhanced: 2.23) and DeepSeek-R1 (base: 3.42; enhanced: 2.37). Qwen-2.5-72B demonstrated notably lower accuracy (base: 5.95; enhanced: 2.75). o3-mini consistently outperformed other models with a significant margin, surpassing GPT-4o (base: *adj. p*lJ<lJ0.001, enhanced: *adj. p*lJ=lJ0.007), DeepSeek-R1 (base: *adj. p*lJ<lJ0.001, enhanced: *adj. p*lJ< 0.001), and Qwen-2.5-72B (base: *adj. p*lJ<lJ0.001, enhanced: *adj. p*lJ< 0.001).

**Figure 2:**
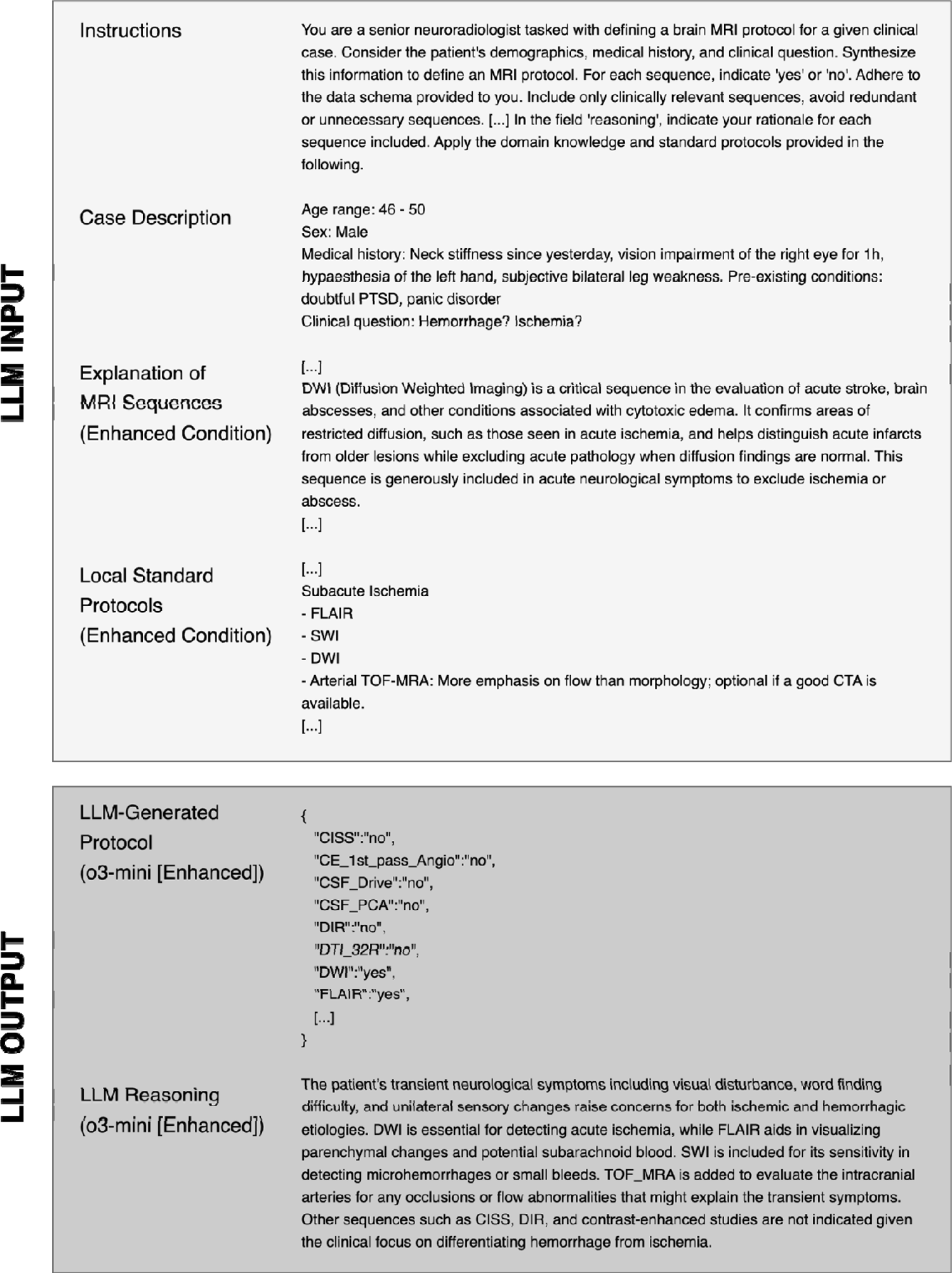
Exemplary brain MRI case. In the enhanced condition, an explanation of MRI sequences as well as local standard protocols were additionally provided to LLMs within the context window. For all models, the structured output mode was applied to obtain responses adhering to a structured JSON (JavaScript Object Notation) schema.

**Figure 3:**
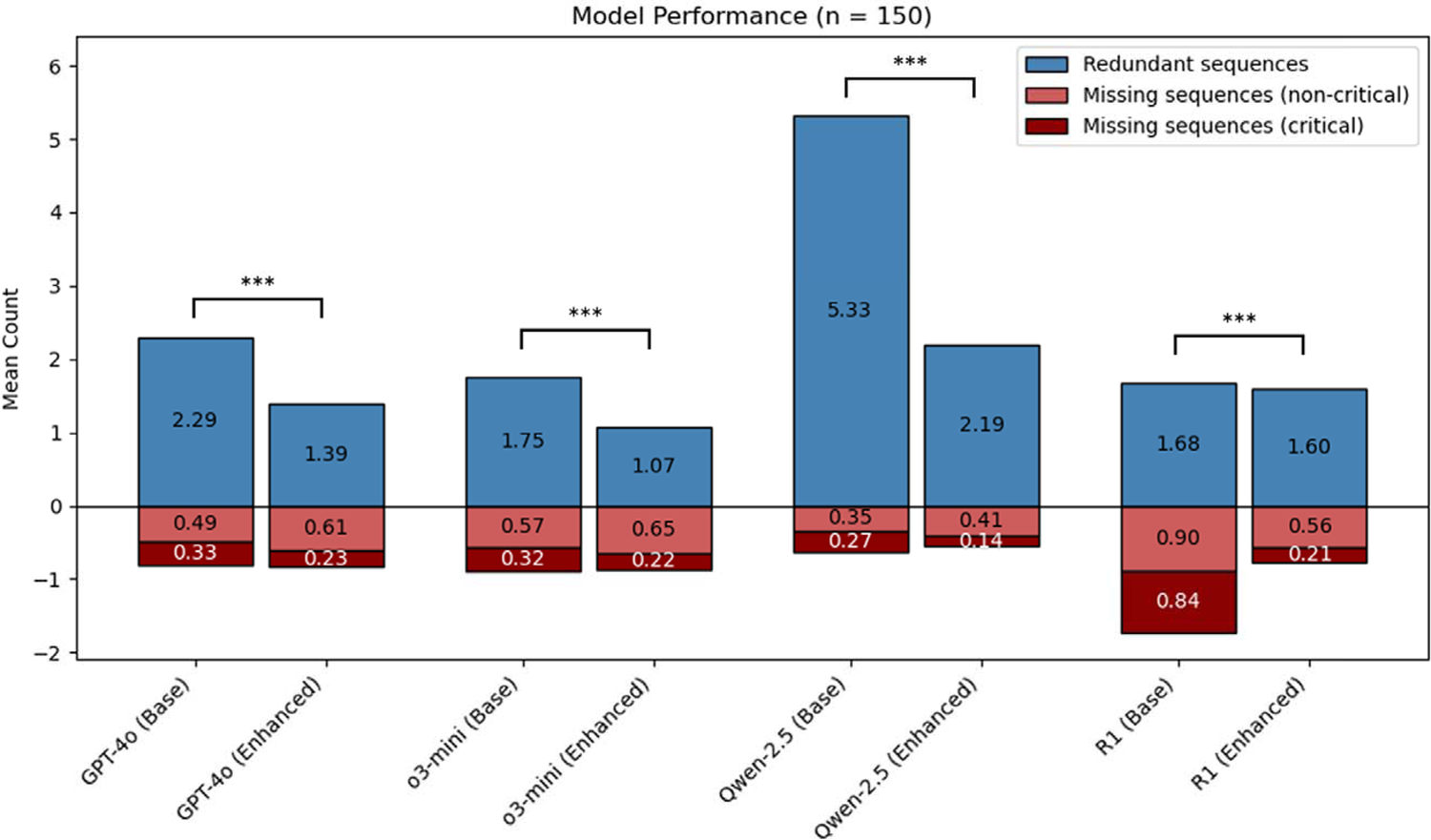
Model performance in defining brain MRI protocols. ‘Enhanced’ indicates that the model received additional domain-specific knowledge - local standard protocols and explanations for the indications of MRI sequences. The accuracy index (sum of missing and redundant sequences) was used to statistically compare models with and without domain knowledge. *** Adj. p < 0.001.

GPT-4o surpassed Qwen-2.5-72B in both the base (*adj.* p < 0.001) and enhanced (*adj.* p < 0.001) conditions, but showed no significant difference compared to DeepSeek-R1 (base: *adj.* p = 0.052; enhanced: *adj.* p = 0.174). All four models showed highly significant improvements in accuracy when augmented with additional domain knowledge (base vs enhanced; *adj. p* < 0.001 for all models). Importantly, this improvement was primarily driven by a substantial reduction of unnecessary (redundant) MRI sequences, whereas the rate of missing critical sequences remained largely stable. For example, enhanced queries with o3-mini reduced redundant sequences from a mean of 1.75 (base) to 1.07 (enhanced), while the frequency of missing sequences remained largely unchanged (base: 0.89; enhanced: 0.87). In all scenarios, except for base queries using DeepSeek-R1, models recommended more redundant than missing sequences, reflecting a general tendency toward overly comprehensive protocol recommendations.

In Qwen-2.5-72B, the enhanced condition led to a major decrease of cases with unnecessary contrast administration (base: 95.3% [143/150], enhanced: 54.0% [81/150], *adj.* p < 0.001), whereas the opposite trend was seen in DeepSeek-R1 (base: 23.3% [35/150], enhanced: 35.3% [53/150], *adj.* p = 0.012) (Figure 4).

**Figure 4:**
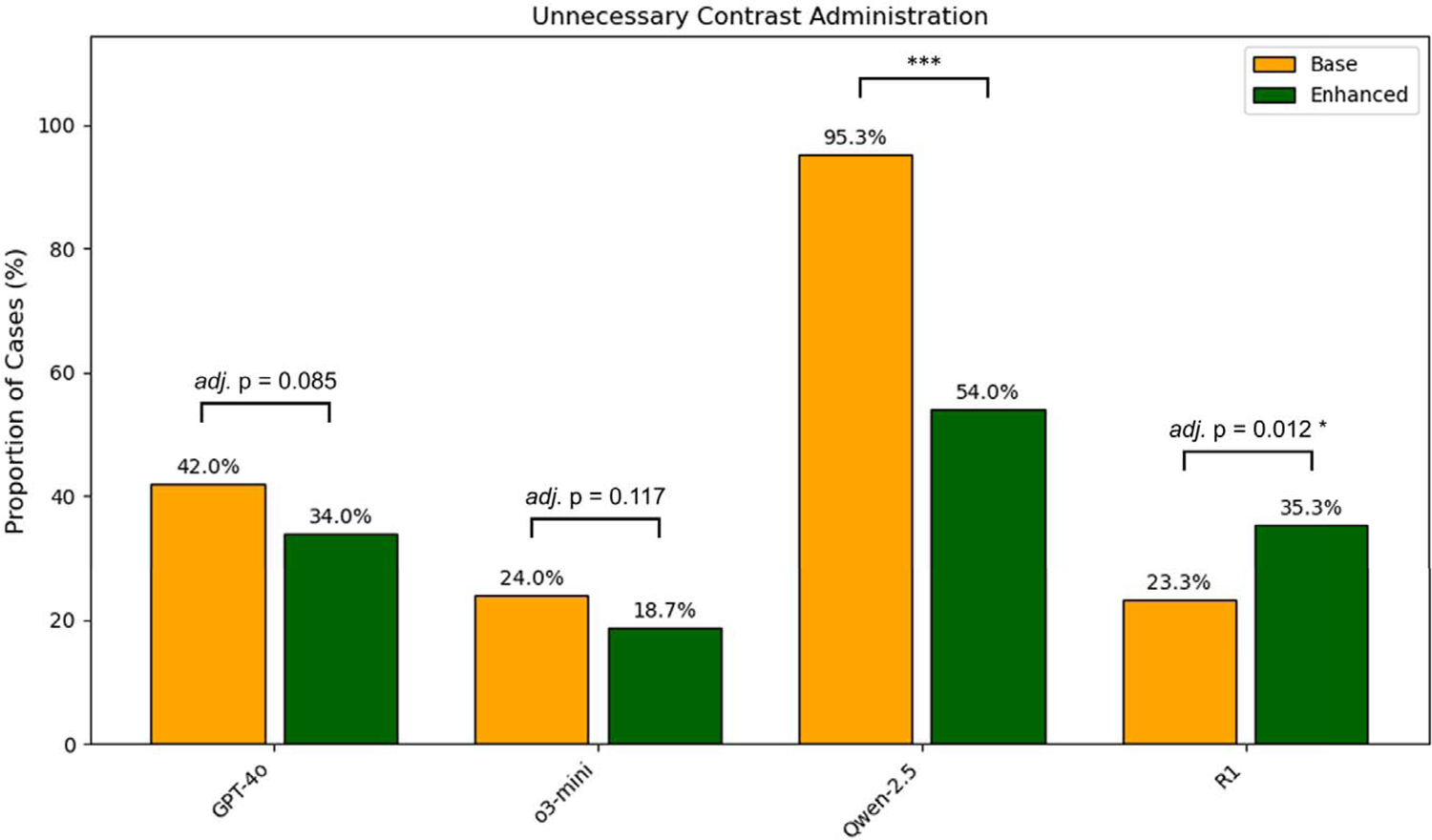
Proportion of cases with LLM-generated protocols suggesting unnecessary contrast administration. ** Adj. p < 0.01. *** Adj. p < 0.001.

Protocoling accuracy varied between case types. Across all four models, the greatest protocoling accuracy was seen for ‘degenerative’ cases (accuracy index [base]: 2.48, accuracy index [enhanced]: 1.73). In contrast, inferior protocoling performance was observed in ‘vascular’ cases (accuracy index [base]: 4.34, accuracy index [enhanced]: 2.47) and ‘miscellaneous’ cases (accuracy index [base]: 4.19, accuracy index [enhanced]: 2.80) (Figure 5).

**Figure 5:**
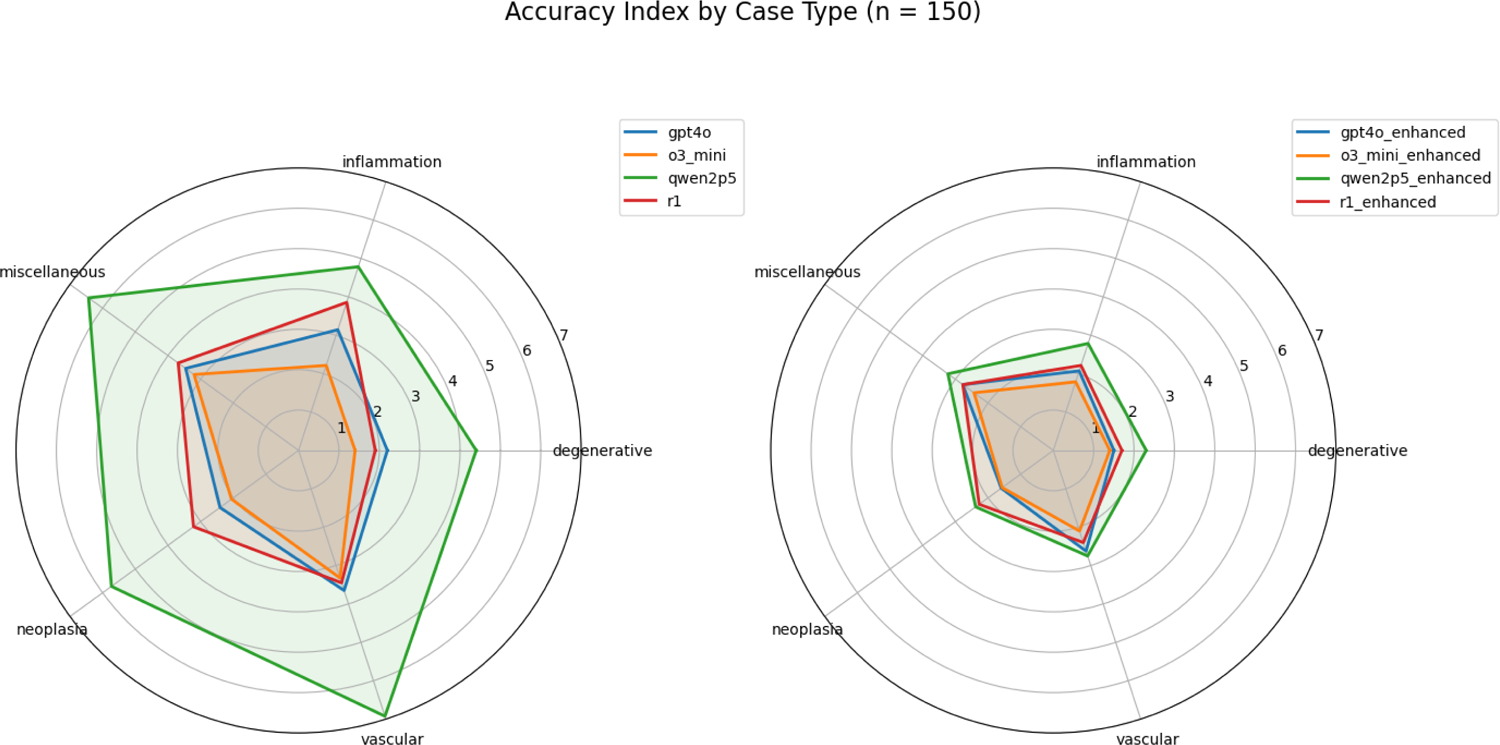
Accuracy index by case type (n = 150). Accuracy index was defined as the sum of missing and redundant sequences in LLM-generated protocols, as compared to the consensus protocol of two board-certified neuroradiologists. Higher accuracy index indicates lower protocoling accuracy.

### Resident Protocoling Performance

In the subsample of 50 cases, the two radiology residents achieved accuracy indices of 1.80 (0.98 redundant and 0.82 missing sequences per case) and 1.44 (0.58 redundant and 0.86 missing sequences per case), respectively. The highest-performing LLM (o3-mini [enhanced]) demonstrated comparable protocoling accuracy, reaching an index of 1.92 (1.16 redundant and 0.76 missing sequences per case) (Figure 6).

**Figure 6:**
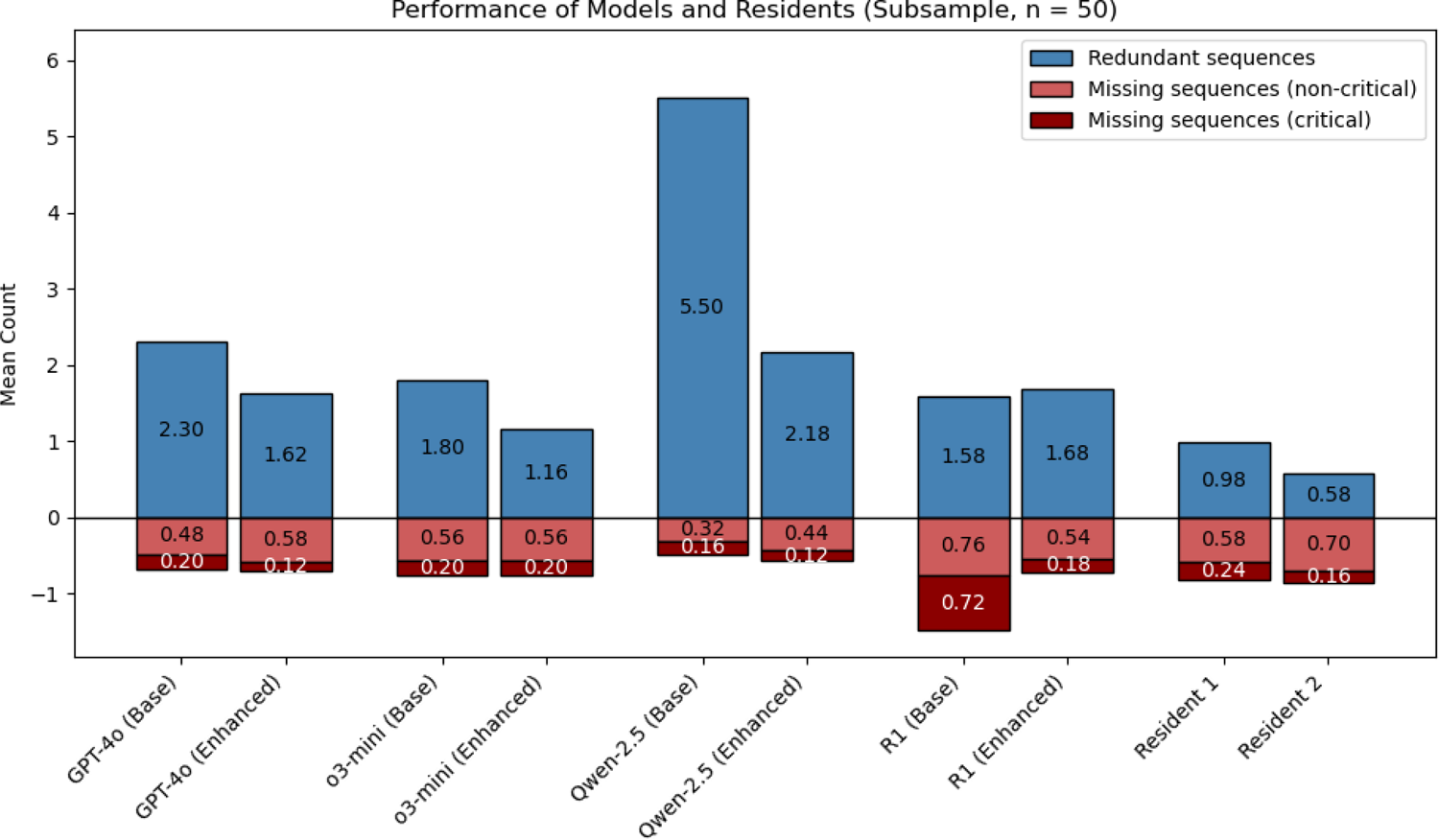
Performance of models and residents in defining brain MRI protocols (subsample, n = 50). ‘Enhanced’ indicates that the model received additional domain-specific knowledge - local standard protocols and explanations for the indications of MRI sequences. Residents 1 and 2 had 18 and 16 months of dedicated neuroradiology experience at the time of the study.

## Discussion

This study evaluated the performance of four state-of-the-art LLMs in creating granular, sequence-level brain MRI protocols with and without augmentation through in-context learning.

In summary, LLMs showed promising performance in brain MRI protocoling. In-context learning involving local standard protocols and explanations of MRI sequences consistently resulted in a substantial and highly significant improvement of protocoling accuracy. Notably, the highest-performing LLM (o3-mini with enhanced queries) yielded accuracy levels comparable to those of radiology residents with around 1.5 years of dedicated neuroradiology experience, demonstrating potential for automating this time-consuming process. Our study extends the findings of previous studies revealing the potential of LLMs in (semi-)automating imaging protocols (11–15). Although fictitious patient cases were used, the generalizability of the findings is unlikely to be affected, as the data was derived from real-world patient records through only minor modifications irrelevant to the MRI protocol.

However, several obstacles remain to be addressed before (semi-)automation of MRI protocoling is attainable. A technical infrastructure ensuring the preservation of patient data privacy – either through deployment on local hardware or a secure cloud-based platform - is prerequisite for any clinical implementation of an LLM-based system (17). Open-weight models such as DeepSeek-R1 - which demonstrated reasonable performance in this study - present a viable option for local deployment. Additionally, LLM-based systems should be optimized and tested for the prevention of any patient harm. Defining an imaging protocol involves the exclusion of potential contraindications for a particular imaging modality (e.g. presence of a pacemaker) or contrast administration (e.g. known allergy) (18). Avoiding any threats arising from complete automation and overreliance on autonomous decision-making systems (a psychological phenomenon also known as automation bias (19,20)) is critical. Insufficient clinical histories in imaging request forms are another frequent challenge faced by radiologists, requiring additional manual review of the electronic medical records (EMR). Recent studies have explored the possibility to utilize LLMs to assess the completeness of clinical histories (21) and augment request forms by mining the EMR for pertinent information (13). In the future, an agentic LLM system capable of orchestrating multiple tasks autonomously (22) could support an end-to-end auto-protocoling workflow, assessing potential contraindications, enriching clinical histories with EMR-extracted data, and eventually generating a tailored imaging protocol.

In this study, LLMs were enhanced using in-context learning, leading to improved protocol accuracy. The enrichment of LLM queries with external information sources has been an area of considerable scientific interest, holding promise for enhanced response accuracy while mitigating hallucinations (23). A widely adopted approach, retrieval-augmented generation (RAG), utilizes a pre-indexed external database to dynamically retrieve relevant information during response generation (24). However, with the advent of LLMs featuring extensive context windows, vast amounts of information can now be processed directly through in-context learning approaches, eliminating the need for sophisticated retrieval architectures – although at the cost of increased computational resources (25). As of this writing, Gemini 2.0 Pro (Google LLC, Mountain View, USA) features the largest context window, accommodating up to 2 million tokens, equivalent to approximately 3,000 pages of text (26). Spanning 4,013 tokens only, the external documents used in this work were sufficiently compact to fit within the context window of all evaluated LLMs. The option to easily replace the external documents may allow for seamless adaptation to institution-specific protocoling standards.

Intriguingly, the MRI protocoling performance of GPT-4o – a non-reasoning LLM – was only slightly inferior to o3-mini, and comparable to DeepSeek-R1, both of which are considered state-of-the-art reasoning LLMs (27,28). Reasoning models, designed to decompose problems into smaller logical steps through techniques like chain-of-thought, are known to excel at complex mathematical or logical reasoning tasks. Compared to non-reasoning LLMs, they have been reported to better handle tasks outside their training data, albeit at the cost of increased processing time (29). In clinical settings, these structured reasoning capabilities could contribute to improved transparency and explainability, allowing clinicians to better understand and validate the model’s output. Although direct comparisons were constrained by the undisclosed sizes of o3-mini and GPT-4o, our results suggest that non-reasoning LLMs can perform competitively across various tasks, sometimes matching reasoning models. This underlines the importance of tailoring model selection to the specific task requirements.

## Limitations

This study has several limitations. First, to ensure feasibility of the analysis, specific MRI protocol parameters that influence exam assessability, such as orientation and slice thickness, were not considered. Second, some MRI sequences (T1 MPRAGE and T1 Dixon, T2* and SWI) were treated as interchangeable, despite their distinct technical properties and slightly different diagnostic utilities in evaluating particular anatomical structures or pathologies. Third, the accuracy index used to evaluate LLM protocoling performance implicitly gave equal weight to redundant and missing sequences, although omission of sequences could have greater clinical significance. Albeit imperfect, this approach allowed the combined evaluation of clinical relevance and operational efficiency in a single metric. Lastly, this is a single-center study and its findings require external validation across multiple institutions, particularly given the variability of available MRI sequences and protocoling standards.

In conclusion, we demonstrate promising potential of LLMs in automating brain MRI protocoling, especially when augmented with local protocoling standards. o3-mini exhibited superior performance, followed by GPT-4o.

## Data Availability

All data produced in the present study are available upon reasonable request to the authors.

