## Supplements for "Evaluating Large Language Model-Generated Brain MRI Protocols: Performance of GPT4o, o3-mini, DeepSeek-R1 and Qwen2.5-72B"

| Case Type | Standard Brain MRI Protocol |
| --- | --- |
| Headache/Sinus Venous Thrombosis (SVT) | - FLAIR: Swelling of inadequately drained parenchyma?  - DWI: Congestive ischemia?  - T2: Transverse sinus. Hyperintense gaps (thrombus)? Hypoplasia?  - SWI: Secondary congestion hemorrhage?  - 4D TRAK: Start 10s after injection. Asymmetric perfusion?  - MPRAGE: After contrast. Caution: Chronic thrombi may be contrast-enhancing and thus appear masked. |
| Dizziness | - FLAIR  - DWI: Brainstem/cerebellar ischemia? If brainstem ischemia suspected.  - Arterial TOF-MRA: Vascular-nerve contact? (Usually AICA loop with cranial nerve VIII, often physiological without pathological relevance)  - T1 Dixon  - CISS: Start 1 min after contrast. Visualization of cranial nerve exit/internal auditory canal.  - T1 Dixon: After contrast. Space-occupying lesion in cerebellopontine angle (schwannoma → contrast-enhancing?). |
| Multiple Sclerosis (MS) | - DIR  - FLAIR  - MPR  - MPR with contrast: Optional after contrast if acute relapse suspected. |
| Optic Neuritis | - DIR  - FLAIR  - MPR  - T1 Dixon: Orbit  - T2 Dixon: After contrast  - MPR with contrast  - T1 Dixon: After contrast, orbit |
| Abscess/Encephalitis | - FLAIR  - DWI  - SWI: Useful for differentiating fungal abscess (central susceptibility) vs. bacterial abscess ('dual-rim sign'). Hemorrhagic encephalitis (e.g., herpes encephalitis) → cortical susceptibility. Rare hemorrhagic encephalitides: Japanese encephalitis/Dengue.  - MPR  - T2 TSE: If encephalitis is suspected, scan hippocampus; otherwise, scan over abscess.  - MPR with contrast  - FLAIR with contrast: Optional. Further delineation of leptomeningeal enhancement → subarachnoid empyema? |
| Meningitis | - FLAIR  - SWI  - T1 MPRAGE  - T1 MPRAGE with contrast  - FLAIR with contrast |
| Vasculitis (Intra- and Extracranial) | - FLAIR: Past ischemia as an indicator of chronic progression?  - SWI: Asymmetric intracranial vessel visualization?  - DWI: Secondary ischemia due to vasoconstriction?  - Arterial TOF-MRA: Look for caliber changes ('string of beads' pattern)?  - 3D T1 SE BB  - 3D T1 SE BB with contrast: Caution: Often 'false-positive' enhancement of carotid branches; compare with the contralateral side. Temporal arteritis cannot be reliably diagnosed by MRI. |
| Subacute Ischemia | - FLAIR  - SWI  - DWI  - Arterial TOF-MRA: More emphasis on flow than morphology; optional if a good CTA is available. |
| Brainstem Ischemia | - FLAIR  - DWI  - SWI  - T2: Focus on brainstem. Should primarily be used to demarcate infarcts, as FLAIR is often artifact-prone near the skull base.  - Arterial TOF-MRA: More emphasis on flow than morphology; optional if a good CTA is available. |
| Dissection | - FLAIR  - SWI  - DWI: Ischemia in the affected vascular territory → acute therapeutic implications (e.g., lysis)  - T1 SPIR: From carotid bifurcation to carotid siphon. Crescent-shaped intraluminal hematoma, hyperintense after approx. 3 days.  - T2 SPAIR: Same as T1 SPIR. Time-dependent signal changes.  - High-resolution neck angio: Bolus triggering. |
| Hypoxic Brain Injury | - FLAIR: Hyperintensity in basal ganglia?  - DWI  - T1 FFE: Hyperintensity in laminar cortical necrosis? Blurred corticomedullary differentiation?  - SWI |
| Glioma | - FLAIR  - MPRAGE  - DWI  - PWI: After contrast  - T2  - MPRAGE with contrast |
| Epilepsy | - FLAIR: Focus search: Mass lesion? FCD: Transmantle sign?  - SWI: Focus search: Hemorrhage?  - DIR: FCD: Transmantle sign? Focus search: Cortical developmental disorders?  - T2: Tilt over hippocampus. Mesial temporal sclerosis → Temporal lobe epilepsy?  - MPRAGE: Volumetric analysis.  - DWI: Only in the acute phase; often diffusion restriction in hippocampus/pulvinar/cortex. |
| Dementia | - FLAIR  - SWI: Microbleeds? Indications for Lewy body disease?  - MPRAGE |
| Parkinson's Disease | - FLAIR  - SWI  - T2  - MPRAGE |
| Encephalopathy | - FLAIR: Wernicke encephalopathy: Hyperintensity in mammillary bodies/dorsomedial thalamus/mammillary bodies. Toxic encephalopathy: Hyperintensities generally corpus callosum-associated and in centrum semiovale. Hepatic encephalopathy: Insular hyperintensities, posterior limb of internal capsule, thalamus. NBIA: 'Eye of the tiger' in anteromedial globus pallidus. Wilson's disease: 'Face of the giant panda' in midbrain.  - MPR  - DWI: Often diffusion restriction overlapping with FLAIR changes.  - SWI: If Wilson's disease or NBIA is suspected (hypointensity of basal ganglia due to copper/iron accumulation).  - T2 with contrast  - MPR with contrast |
| Cranial Nerves | - FLAIR  - T1 Dixon  - 3D CISS: After contrast  - TOF: Optional if vascular-nerve contact is suspected.  - DWI: Optional if acute pathology is suspected.  - T1 Dixon: After contrast |
| Normal Pressure Hydrocephalus | - T2 FLAIR: Callosal angle, DESH, CSF leakage, etc.  - CSF Drive: Aqueduct/foramina of Monro.  - CSF PCA: Flow visualization.  - T2: Aqueduct stenosis? |
| Pituitary Mass | - FLAIR  - T2  - T1: Parallel to the pituitary stalk.  - T1 dynamic: Not for follow-up! Only for initial detection, as pituitary adenomas typically enhance slowly. Start contrast and scan simultaneously.  - T1 after contrast |
| Orbit | - FLAIR  - T1 Dixon: Orbit  - T2 Dixon  - T1 Dixon: After contrast, orbit |

Supplement 1: Local standard brain MRI protocols.

| MRI Sequence | Explanation |
| --- | --- |
| CISS | CISS (Constructive Interference in Steady State) is a high-resolution imaging sequence used to visualize cranial nerve anatomy, inner ear structures, and cisternal spaces. It is particularly valuable in evaluating lesions in the cerebellopontine angle, such as vestibular schwannomas, and in diagnosing neurovascular compression syndromes. This sequence confirms small lesions, cystic changes, or abnormal cerebrospinal fluid (CSF) relationships around cranial nerves while excluding subtle anatomic abnormalities that might be missed on conventional imaging. However, if the clinical question does not involve the cranial nerves, inner ear, or detailed CSF anatomy, this sequence can be omitted to reduce scan time. |
| CE 1st pass angiography | CE_1st_pass_Angio (Contrast-Enhanced First-Pass Angiography) captures the first pass of contrast through the cerebral vasculature, which is particularly valuable for the assessment of potential reoccurences after endovascular treatments of primarily aneurysms. This sequence is only necessary after stenting or flow diverter implantation. |
| CSF Drive | CSF_Drive is useful in assessing CSF flow dynamics when there is a suspicion of subtle distortions in CSF pathways due to cysts, small masses, or adhesions. The sequence confirms normal CSF anatomy and can detect minor alterations or obstructions, effectively excluding lesions that might distort the CSF spaces. When the clinical focus is not on CSF dynamics or anatomy, and other sequences already provide adequate CSF detail, this sequence is not necessary. |
| CSF PCA | CSF_PCA is indicated when there is a need to evaluate the dynamics of CSF flow, such as in cases of hydrocephalus, Chiari malformations, or suspected CSF leaks. By quantifying flow velocities and directions, it confirms abnormal flow patterns and excludes functional blockages if normal flow is observed. In patients where CSF flow disturbances are not suspected, or when anatomical sequences are sufficient, this sequence is not necessary. |
| DIR | DIR (Double Inversion Recovery) is particularly useful for detecting cortical and juxtacortical lesions, making it a valuable tool in the assessment of multiple sclerosis. By simultaneously suppressing the signals from both CSF and white matter, DIR confirms subtle cortical lesions that might otherwise be overlooked and excludes minor cortical abnormalities in routine imaging where such detailed evaluation is not required. |
| DTI | DTI_32R (Diffusion Tensor Imaging with 32 Directions) provides an assessment of white matter integrity and is instrumental in performing fiber tractography. This sequence is only necessary in preoperative imaging for navigated surgery, as explicitly indicated in the clinical question. |
| DWI | DWI (Diffusion Weighted Imaging) is a critical sequence in the evaluation of acute stroke, brain abscesses, and other conditions associated with cytotoxic edema. It confirms areas of restricted diffusion, such as those seen in acute ischemia, and helps distinguish acute infarcts from older lesions while excluding acute pathology when diffusion findings are normal. This sequence is generously included in acute neurological symptoms to exclude ischemia or abscess. |
| FLAIR | FLAIR (Fluid Attenuated Inversion Recovery) is essential for detecting lesions near CSF spaces, particularly in conditions like multiple sclerosis, infections, or subarachnoid hemorrhage. By suppressing the bright CSF signal, FLAIR confirms parenchymal abnormalities such as demyelinating plaques, edema, or gliosis, and it effectively excludes subtle white matter changes when the signal is normalized. Given its diagnostic value, FLAIR is included in all brain MRI studies. |
| FLAIR post-contrast | FLAIR_post_contrast is used when there is a need to identify subtle leptomeningeal or parenchymal enhancement that might not be apparent on standard T1 post-contrast images. This sequence confirms pathological enhancement in areas where CSF signal suppression is critical, thereby excluding inflammatory or infectious processes if no enhancement is observed. It is only necessary to exclude encephalitis or meningitis. |
| Neck angiography | Neck_angiography targets the cervical vasculature and is indicated when there is a suspicion of carotid or vertebral artery pathology, such as dissection or stenosis. This sequence is only necessary when a cervical artery dissection needs to be excluded or in primary MRI imaging in acute stroke. |
| T1 Dixon | T1_Dixon is a technique used when robust fat–water separation is required to better characterize tissue composition. It is particularly useful in regions where fat signal might mimic or obscure pathology, such as at the skull base or within the orbits, by confirming the presence of fat versus other tissue components and excluding misinterpretations of fat as pathology. |
| T1 Dixon post-contrast | T1_Dixon_post_contrast builds on the benefits of the Dixon technique by combining it with contrast enhancement, making it especially useful in areas with abundant fatty tissue. It confirms the extent and nature of contrast enhancement while avoiding artifacts related to fat signal, thus excluding false positives due to unsuppressed fat. If standard post-contrast T1 images provide sufficient contrast, the addition of this sequence may not be necessary. |
| T1 MPRAGE | T1_MPRAGE (Magnetization Prepared Rapid Gradient Echo) is a high-resolution, three-dimensional anatomical sequence. It confirms subtle anatomical abnormalities and small lesions with high spatial resolution, thereby excluding pathology when the brain structure appears normal. Among other cases, this sequence is necessary to visualize intraparenchymal supratentorial lesions, along with T1_MPRAGE_post_contrast. In addition, this sequence is necessary when corticomedullary differentiation is important to visualize (MS, dementia, parkinson, epilepsy). |
| T1 MPRAGE post-contrast | T1_MPRAGE_post_contrast is employed to detect lesions that enhance following contrast administration, which is vital for identifying tumors, infections, or areas of inflammation. This sequence confirms blood–brain barrier breakdown by demonstrating contrast uptake, and it excludes non-enhancing lesions when normal contrast distribution is observed. It is invariably acquired along with T1_MPRAGE (pre-contrast). |
| PWI | PWI (Perfusion Weighted Imaging) is only necessary in glioma imaging. It confirms areas of altered perfusion, helping to assess tumor vascularity, and it excludes perfusion abnormalities when normal blood flow is observed. |
| SWI | SWI (Susceptibility Weighted Imaging) is a highly sensitive sequence for detecting microhemorrhages, calcifications, venous anomalies, or iron deposition. It confirms the presence of blood products or calcifications that might be invisible on other imaging sequences, and it excludes hemorrhagic or calcific pathology when the SWI signal is unremarkable. |
| T1 BB | T1_BB (T1 Black Blood) is designed for vessel wall imaging by suppressing the signal from flowing blood. This sequence is essential for assessing vasculitis by comparing the native imaging with post contrast imaging and assessing for vessel wall enhancement. This sequence is only required for the diagnosis of vasculitis. |
| T1 BB post-contrast | T1_BB_post_contrast in used for vessel wall imaging by combining the black blood technique (suppression of blood signal) with contrast medium administration. In inflammatory or neoplastic involvement, vessel walls could show enhancement. It excludes significant vessel wall disease when no enhancement is present. Aside from this, the BB post contrast should always be included when examining cerebral metastases due to its excellent sensitivity. In the assessment of cerebral metastases, only the BB post contrast and no native BB is required. |
| T1 SPIR | T1_SPIR (Spectral Presaturation with Inversion Recovery) is used to achieve effective fat suppression in T1-weighted imaging, particularly in post-contrast studies. It is only used in the assessment of cervical artery dissections. |
| T1 dynamic contrast-enhanced | T1_dynamic_contrast_enhanced imaging is only used in the initial diagnostic assessment of (suspected) pituitary microadenomas. |
| T2 | T2 imaging can be useful for assessing fluid content, edema, cysts, and other pathology such as gliosis or demyelination. In practice however, T2 FLAIR serves the same purpose. Thus, when T2 FLAIR is already in a protocol, T2 is not essential. However, when assessing pathologies in and around the brain stem, the sequence needs to be included. |
| T2 Dixon | T2_Dixon is used when fat–water separation on T2 images is necessary, especially in regions adjacent to fatty tissue. By clearly delineating fat from edema, it confirms the nature of tissue components and excludes misinterpretation of fat as an abnormal signal. This sequence is used only in the context of orbital pathologies, such as when the optic nerve or orbital muscles are of primary interest |
| T2 SPAIR | T2_SPAIR (Spectral Attenuated Inversion Recovery) offers robust fat suppression on T2 images, thereby enhancing the conspicuity of lesions in areas where fat might otherwise obscure pathology. ItIt is only necessary for the exclusion of cervical artery dissection. |
| TOF-MRA | TOF_MRA (Time-of-Flight Magnetic Resonance Angiography) is a non-contrast, static technique used to image the intracranial arteries. It confirms the patency and course of major vessels by demonstrating normal flow, and it excludes significant stenoses or occlusions when normal flow patterns are observed. TOF-MRA a basic sequence for static vessel imaging and should be included in protocols that aim to assess cerebral arteries. |
| TRAK | TRAK (4D) is a time-resolved angiographic sequence requiring contrast medium, which offers dynamic imaging of blood flow patterns, making it particularly useful in evaluating arteriovenous malformations or vascular shunts. By providing real-time flow information, it confirms abnormal flow dynamics and excludes static vascular anomalies. This sequence is only necessary when dural arteriovenous fistulae, arteriovenous malformations, or cerebral sinus venous thrombosis need to be excluded. |
| TRANCE | TRANCE (4D, Triggered Angiography Non-Contrast Enhanced) is a time-resolved, non-contrast, flow-sensitive angiographic technique that dynamically images vascular structures. Its dynamic nature confirms normal versus abnormal flow patterns, including conditions of either reduced or heightened flow. This is particularly useful when assessing conditions such as arteriovenous malformations or dural arteriovenous fistulas. If other non-contrast techniques such as TOF-MRA already yield the necessary vascular details, TRANCE is not necessary. |
| TOF-MRA post-contrast | TOF_MRA_post_contrast combines the benefits of time-of-flight imaging with contrast enhancement to achieve even better delineation of the vascular lumen. This sequence confirms subtle vascular irregularities by increasing the signal-to-noise ratio in regions of slow or turbulent flow and excludes false negatives that might occur with non-contrast methods. This sequence is only necessary following stenting or flow diverter implantation. |

Supplement 2: Explanation of MRI sequences.
